# Intra-Arrest Percutaneous Stellate Ganglion Block: A Protocol for a Scoping Review

**DOI:** 10.1101/2024.05.30.24308206

**Authors:** Ruan Vlok, Ravi Shankar, Jamie Cham, Bradley Bridge, Lachlan Donaldson

## Abstract

**Introduction:** Percutaneous Stellate ganglion blockade (PSGB) is an increasingly recognised technique for the management of refractory ventricular arrhythmias (VA). Both ultrasound guided and landmark based techniques have been used to successfully decrease the burden of VA. The role of intra-arrest PSGB remains unknown, however it may represent a cost-effective point of care intervention that can be performed for shockable cardiac arrest.

**Methods:** A systematic review of all clinical studies of PSGB performed intra-arrest will be performed across multiple databases. Studies will be included if they described the use of intra-arrest PSGB in humans. Studies will be qualitatively assessed to describe data regarding the PSGB technique, the training of the proceduralist, patient demographics, the context of the arrest and clinical outcomes and complications. This protocol was drafted according to the PRIMSA-P ScR.

## Introduction

Percutaneous Stellate Ganglion Block (PSGB) has been widely described as an intervention for a variety of clinical conditions, including increasing recent interest in its role in refractory ventricular arrhythmias (VA)^1^. The recently published STAR study demonstrated that the procedure can be performed safely and reported effective VA suppression in 92% of patients with electrical storm^2^. This procedure therefore represents a potential intervention for patients in refractory cardiac arrest in shockable arrhythmias.

Refractory shockable cardiac arrest is a pathology with increasing treatment complexity. The 2023 AHA focussed update on advanced cardiac life support (ACLS) gave ECPR a grade 2A recommendation, a higher recommendation than amiodarone and adrenaline^3^. However, ECPR is a cost and resource intensive intervention that is not universally available. The recent DOSE-VF Trial reported increased rates of return of spontaneous circulation (ROSC) with vector change or double sequential external defibrillation (DSED), a simple intervention which represents an opportunity to intervene earlier in the cardiac arrest^4^. Likewise, PSGB conceivably represents a cost effective and simple intervention that can be performed as an adjunct to ECPR, or when ECPR is unavailable.

PSGB can be performed by landmark, ultrasound or fluoroscopy guidance. It is typically performed as a unilateral left sided procedure for VA suppression, although bilateral procedures are described^2^. Both single shot and continuous infusion studies have been described. Concerns for adverse events include vascular injury, intravascular infiltration of local anaesthetic (LA), hoarseness, phrenic nerve palsy and upper limb palsy. The START study reported 67% of procedures were performed in patients on patients on dual antiplatelets and/or anticoagulation and described a single major complication^2^. This complication was an episode of systemic local anaesthetic uptake in a patient already on a lignocaine infusion.

This safety data appears in keeping with data assessing the anatomical approach to PSGB which described no significant complications in approximately 2000 patients receiving the block for analgesic purposes^5^. Performance of the procedure mid-cardiac arrest represents suboptimal conditions, and it is important to determine the safety profile of the procedure in this context before widespread uptake can be recommended. The procedure also represents a potential distraction of proven resuscitation priorities.

We will aim to perform a scoping review of reported literature of PSGB performed during cardiac arrest to describe the state of the literature.

## Methodology

A scoping review of the use of intra-arrest PSGB will be performed to provide a narrative overview of the available literature. A single specific question will not be sought to be answered, but a description of the clinical context of published cases and their reported outcomes will be compiled. This protocol will be compiled according to the Preferred Reporting Items for Systematic Reviews and Meta-analysis Protocols for Scoping Reviews (PRISMA-P ScR)^6^.

### Database Search

We will perform a comprehensive electronic database search for articles using key words, synonyms and subject headings that relate to ventricular arrhythmias, cardiac arrest and stellate ganglion block. We will use controlled vocabulary specific to each database (see ‘Appendix 1’). Two review authors will perform the search independently.

We will search the following databases for published studies:

1. Cochrane Central Register of Controlled Trials (CENTRAL) (1996 to present)
2. MEDLINE Ovid (1946 to present)
3. Elsevier Embase (1947 to present)
4. CINAHL EBSCO (1937 to present)

We will perform a citation search of all included studies as well as any relevant studies and reviews on PSGB for in cardiac arrest. We will search the grey literature search using google scholar.

### Criteria for Inclusion

Studies will be included if they report human cases of the use of PSGB in cardiac arrest. Case reports, case series, retrospective and prospective observational studies and randomised controlled trials (RCT) will be eligible. Conference abstracts will be considered eligible for inclusion. Animal studies, reports in healthy volunteers and studies reporting only on the use of PSGB in refractory ventricular arrhythmias with a pulse will be excluded.

### Data Collection and Analysis

Two review authors will independently screen all titles and abstracts of each reference identified by our search and independently assessed the full text of any potentially relevant studies for eligibility or exclusion. Covidence software^7^ will used to collate search results, remove duplicates and record screening decisions and exclusions at each stage. Any disagreements will be resolved by discussion and consensus prior to proceeding at each stage.

Through Covidence, a standardized digital data-extraction sheet will be used. Two review authors independently will extract information regarding study design, data regarding the patient demographics, cardiac arrest characteristics, PSGB technique and outcomes. Where required, individual trial authors or organizations were contacted to obtain missing data or clarification regarding unclear data. Any disagreements will be resolved by discussion and consensus. All studies will be assessed for country of origin, authors, primary institution, and years of data collection to ensure outcomes were not collected from overlapping cohorts.

Data collected will include:

1. Study characteristics
  - Methodology
  - Sample size
  - Country of origin
2. PSGB technique data
  - Primary specialty of the proceduralist
  - Previous training in PSGB
  - Ultrasound guided vs landmark technique
  - Unilateral vs bilateral PSGB
  - Single shot vs continuous infusion
  - Pause in CPR?
  - Local anaesthetic formulation and dose
  - Location PSGB performed (prehospital, emergency department, cath lab, operating theatre, intensive care, other)
3. Patient data
  - Age
  - Sex
4. Cardiac arrest related data
  - Presumed cause of arrest
  - Time from arrest to PSGB
  - Time from SGB to defibrillation
  - Number of antiarrhythmics prior to PSGB
  - Survival to hospital admission and discharge
  - Was ECPR employed
  - If ECPR was employed, was the block performed before or after flows were established?
5. Complication related data
  - Haematoma
  - Intravascular injection
  - Pneumothorax
  - Permanent upper limb palsy

Collected data will be tabulated and described narratively.

## Data Availability

All data produced in the present work are contained in the manuscript

## Appendix 1

Search Strategy (OVID layout):

1. Cardiac arrest.mp. or ACLS.tw or CPR.tw or OHCA.tw or heart arrest/
2. (Ventricular arrhythmia or ventricular tachycardia or ventricular fibrillation).tw.
3. heart ventricle arrhythmia/
4. shockable.tw.
5. 1 or 2 or 3 or 4
6. stellate ganglion block/
7. neuromodulation.tw.
8. stellate ganglion.tw.
9. 6 or 7 or 8
10. 5 and 9

